# LLM4TOP: An End-to-End framework based on Large Language Model for trial outcome prediction

**DOI:** 10.1101/2025.07.06.25330956

**Authors:** Long Qian, Xin Lu, Parvez Haris, Li Shuo, Jianyong Zhu, Yingjie Yang

## Abstract

Clinical trial outcome prediction involves estimating the probability of a trial successfully achieving its predefined endpoints. Current approaches primarily employ machine learning techniques that integrate diverse data modalities, including trial protocol descriptions, molecular structures of investigational drugs, and characteristics of target diseases. However, this field faces several critical challenges that hinder practical implementation. The preprocessing of heterogeneous clinical trial data requires extensive and complex transformation pipelines. Different data modalities demand specialized modeling architectures, complicating the development of unified prediction systems. Furthermore, the absence of privacy-preserving large language models capable of local deployment presents a significant barrier to clinical adoption, particularly given the sensitive nature of medical data. Addressing these challenges is essential for advancing reliable and clinically applicable prediction models.

In the present study, we propose llm4top(large language model for trial outcome prediction) which is a framework designed specifically to streamline and standardize the process of Clinical trial outcome prediction using large language model based on Qwen3 [1]. By redefining the task of predicting clinical trial outcomes as a binary classification issue, the complete capabilities of large language models with 8B parameters are efficiently utilized. Findings reveal that the method attains a substantially higher degree of accuracy and precision in predicting clinical trial outcomes. The research underscores the automating and simplifies the complex process work and potential of large language models to significantly enhance the predictive accuracy of clinical trial outcomes, thereby facilitating more effective research efforts, drug development processes, and patient care strategies.

## I. INTRODUCTION

Clinical trials play a vital role in the advancement of new treatments. Drugs need to meet safety and efficacy standards in at least three trial phases before they can be authorized for manufacturing. Carrying out these trials is a time-consuming and expensive undertaking. On average, it takes approximately 10 to 15 years and costs $2.87 billion for a drug to go through all trial phases, and many drugs fail in clinical trials [2]. The digitization of clinical trial records and their outcomes in the past decade has provided an opportunity for more data-driven understandings, which have the potential to enhance the success rate of trials.

Predicting the outcomes of clinical trials is a critical task aimed at estimating the probability of a trial successfully reaching its predetermined endpoints. The utilization of Large Language Models (LLMs), like ChatGPT, has gained extensive recognition in diverse domains such as conversational agents, creative writing, and programming. However, the potential use of LLMs in the medical domain, particularly in forecasting clinical trial outcomes, is a relatively unexplored area. This gap in research presents an opportunity for further investigation into how LLMs can be harnessed to enhance decision-making processes in clinical research.

In the early stages, efforts often focused on predicting individual elements in clinical trials to improve the outcomes of each trial [3][4]. Recently, researchers have started to put forward general methods for trial outcome predictions. For example, Lo et al. [5] predicted drug approvals for 15 disease groups based on drug and clinical trial characteristics by using classical machine learning approaches. Fu et al. [6] recently proposed making use of data from multiple sources (such as molecule information, trial documents, disease knowledge graph, etc.) with an interaction network to capture their correlations and assist in trial outcome predictions.

LLMs undergo an initial pre-training phase using vast amounts of generic data, such as BERT’s [7] pre-training on Wikipedia and Google’s BooksCorpus. This extensive training makes the models highly robust but also renders them universal and not tailored to any particular domain. To align the models with specific use cases and enhance results, additional training or “fine-tuning” is essential. Prominent pre-trained LLMs can be fine-tuned for specific tasks, like text classification.

A major challenge in clinical research is that trial documents are frequently over 1,000 words. Merely encoding long trials by truncating and averaging the embeddings of all remaining tokens will surely result in poor learning quality. Trial2Vec [8] endeavors to address the long-document issue by encoding various sections of the document independently.

Fine-tuning a Large Language Model (LLM) involves using domain-specific text examples as supplementary training data. In the medical domain, using a fine-tuned LLM can significantly improve NLP task performance by adapting the model to the distinct characteristics and subtleties of medical textual data before classification. While non-fine-tuned LLMs may deliver acceptable performance, exploring the use of fine-tuned LLMs, such as llm4top, offers a promising opportunity to further enhance performance in this specific NLP application.

The primary contribution of this study is the innovative application of a state-of-the-art large language model to the domain of clinical trial outcome prediction.

1. This approach shows that treating all input data as text in large language models achieves state-of-the-art performance while bypassing the need for modality-specific architectures (e.g., GNNs for molecular graphs or NLP pipelines for clinical notes), ensuring extensibility without complex preprocessing or dedicated feature extraction.

2. This approach, as far as current knowledge extends, represents the first instance of finetuning a limited sized large language model for the purpose of predicting clinical trial outcomes to be able to keep data privacy without uploading the data through internet.

3. We put forward a novel methodology that utilizes prompt engineering to summarize the long trials without additional training by a large language model.

4. The llm4top framework only needs users to upload an xml file to get the results significantly reduced the learning curve for users and boosted the efficiency of data processing, model selection and result evaluation (see Fig 1.).

**Fig. 1.**
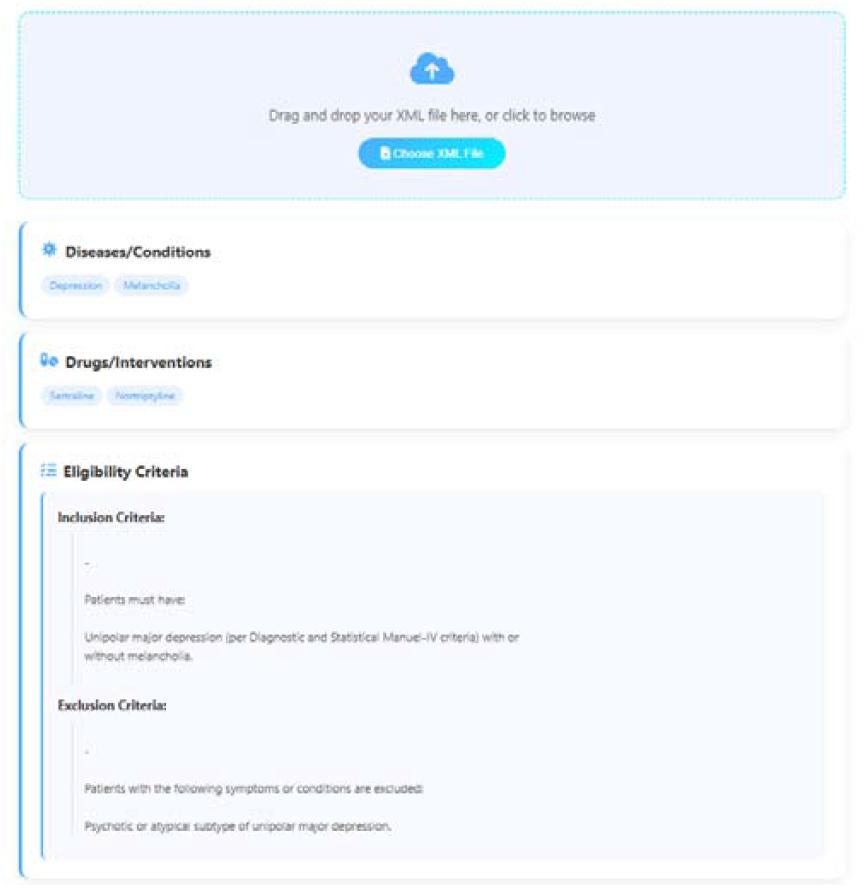
The figure shows a web-based system for analyzing clinical trial data from ClinicalTrials.gov XML files. Users can upload XML files to extract diseases, drug interventions, and eligibility criteria, which are then processed to predict trial success or failure outcomes through an intuitive graphical interface.

## II. RELATED WORK

Previous studies have applied large language models to clinical trial prediction tasks with promising results. For instance, Reinisch et al. adopted LLMs for predicting clinical trial phase transitions [9]. Lai et al. assessed the risk of bias in randomized clinical trials by using LLMs [10]. Jin et al. utilized LLMs to match patients to appropriate clinical trials [11], and Markey et al. leveraged LLMs to assist in the composition of clinical trial documents [12]. Jin et al. conducted an evaluation of LLMs including GPT-4o [13], GPT-3.5 [14], GPT-4 Mini [15], and Llama3 [16] for clinical trial outcome prediction, revealing that these models demonstrated particularly limited efficacy in identifying negative clinical trial outcomes [17]. It shows that it is necessary to retrain the LLMs in medical specific tasks especially clinical trial outcome tasks.

## III. METHOD

The standard prompt-based in-context learning paradigm is like this: when an input sequence x_input_ = {x_1_, x_2_, …, x_n_} is given, the task of assigning a text-class label to an input text is changed into generating a pre-defined textual response y ∈{success, failure} based on the prompt through a language model. The trial outcome prediction task was transformed into a binary classification problem by classifying positive outcomes (for instance, treatment success, disease remission) as one class and negative outcomes (for example, treatment failure, disease progression) as the other.

Since the trial documents are lengthy, typically exceeding 1000 words, we construct a prompt to extract the key information:

> “***Please summarize the following clinical trial inclusion/exclusion criteria to within 1000 characters, retaining key information:***
>
> ***Original text:***
>
> ***{clean_text}***
>
> ***Requirements***
>
> ***1. Retain all important medical conditions and restrictions***
>
> ***2. Maintain the original meaning***
>
> ***3. Remove redundant expressions***
>
> ***4. Keep within 1000 characters***
>
> ***5. Use concise and clear expressions***
>
> ***Summarized criteria:***”

For the zero-shot learning task, the prompt is built based on the following paradigm:

“You are an experienced clinical trial expert. Based on the following clinical trial information, predict the outcome of this trial. Please carefully analyse the disease characteristics, drug mechanisms, and trial protocol. Consider the efficacy, safety, and compatibility of the drug with the target disease. Based on your professional judgment, will this clinical trial be successful? Clinical trial information: ***diseases: {diseases}, drugs: {drugs}, trial protocol: {criteria}. Please only respond with a number 1 (success) or 0 (failure). Do not provide any explanation***.” The reason we add the last sentence is to avoid the large language model output redundant information.

For the Supervised Fine-Tuning task, the prompt is built based on the following paradigm:

***Please predict the outcome of the clinical trial according to the given disease, drugs and trial protocol, output 1 (success) or 0 (failure), the following is the disease, drugs and trial protocol --> diseases: {diseases}, drugs: {drugs}, trial protocol: {criteria}***.

The diseases, drugs and trial protocols are extracted from the TOP clinical trial outcome prediction benchmark from Fu et al [6].

Specific prompt serves two purposes:

(1) providing LLM with evidence to consult on for decision making, which will significantly boost performance.

(2) provides an output format that the outputs of LLM need to follow, so that the output, which is in the form of natural language, can be more easily transformed into labels.

The final dataset for LLM consists of a sequence of annotated examples:

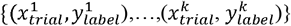

where 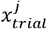, 1 ≤ j ≤ k denotes the j_th_ input sequence and 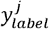 denotes the label, success or failure for the trial outcome prediction task.

The Qwen3-8B **Error! Reference source not found**. is chosen as the base model and the model is finetuned with the Low-Rank Adaptation (lora) method [22], where lora enables parameter-efficient adaptation through low-rank matrix decomposition while preserving the base model’s capabilities. The trained model was to predict the outcomes of future clinical trials which can be formulated as:

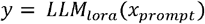

The flow is shown in Fig. 2.

**Fig. 2.**
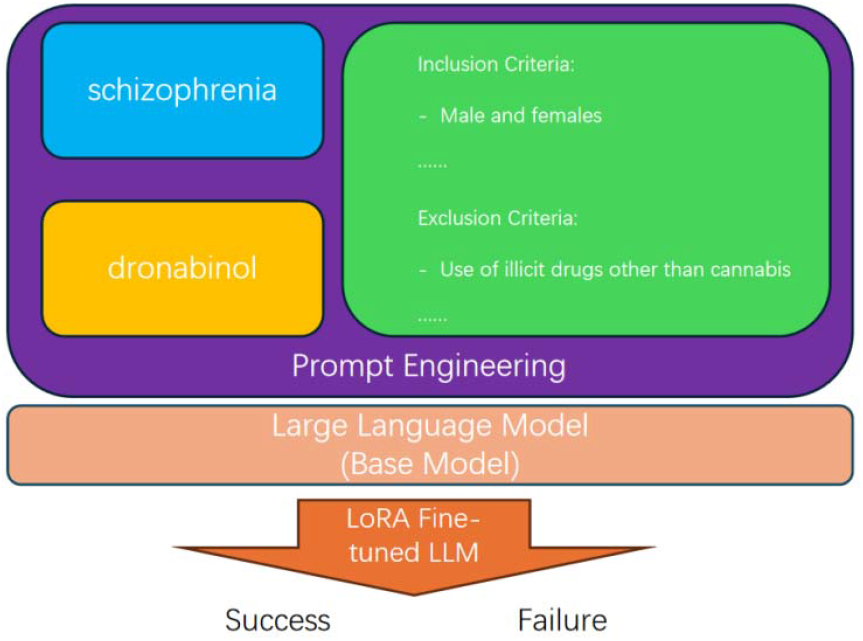
The workflow begins with input data comprising the medical condition (schizophrenia), therapeutic intervention (dronabinol), and inclusion/exclusion criteria. These inputs undergo prompt engineering to structure the data for model processing. A base Large Language Model (LLM) is then fine-tuned using Low-Rank Adaptation (LoRA) methodology, The fine-tuned model processes the structured prompt to generate binary classification outcomes: “Success” or “Failure”

## IV. EXPERIMENT

Hyperparameter Settings. The models are trained for a total of 3 epochs using a mini-batch size of 1 on one NVIDIA 3090 GPU and trained in bf16 precision, which will take up to 2 hours. We employ the AdamW optimizer with a learning rate of 1 × 10^−4^, f3 values of (0.9, 0.99), and a weight decay of 1 × 10^−2^ with a CosineAnnealing learning rate scheduler. The lora rank is 8 and the scale factor is 16.

### Dataset

The TOP clinical trial outcome prediction benchmark proposed by Fu et al. [6] was employed. This dataset contains information regarding drugs, diseases, and eligibility criteria for a total of 17,538 clinical trials. These trials are classified into three phases: Phase I (1,787 trials), Phase II (6,102 trials), and Phase III (4,576 trials). The success rates in dataset differ among phases, with 56.3% of Phase I trials, 49.8% of Phase II trials, and 67.8% of Phase III trials being successful.

Experiments were carried out on different phases of trials separately.

### Metrics

The following performance metrics were considered:

- AUROC: The area beneath the Receiver Operating Characteristic curve, which summarizes the balance between the true positive rate (TPR) and the false positive rate (FPR) across diverse thresholds of FPR. In theory, it is tantamount to computing the ranking quality through the model predictions for identifying the true positive samples. Nevertheless, a higher AUROC does not inevitably imply better-calibrated probability predictions.
- PRAUC: The area beneath the Precision-Recall curve, which summarizes the balance between precision and recall across diverse thresholds of recall. It is equivalent to the average of precision values computed for each recall threshold and is more sensitive to the quality of detecting true positives from the data, such as determining which trial is likely to be successful.
- F1-score: The harmonic mean of precision and recall, taking into account whether the predicted probabilities are well-calibrated, similar to PRAUC.

### Baseline

llm4top was compared against various machine learning and deep learning models, primarily following the setups in [6]:

- Logistic Regression (LR) [18]: This model employs the standard hyperparameters as provided by the scikit-learn library to classify outcomes using logistic regression.
- Random Forests (RF) [18]: We utilize the random forests algorithm, as implemented in scikit-learn, for classification tasks.
- XGBoost [18]: This approach involves the application of a gradient-boosting framework to decision tree algorithms.
- AdaBoost [18]: The AdaBoost method, available through scikit-learn, leverages adaptive boosting to enhance the performance of decision trees.
- kNN+RF [18]: To address missing data, this method first employs k-nearest neighbors for imputation before applying random forests for predictive modeling.
- Feedforward Neural Network (FFNN) [19]: Our FFNN architecture mirrors the feature set of reference [6] and consists of three dense layers with dimensions corresponding to the input feature size, 500, and 100 neurons, respectively, utilizing ReLU activation functions.

For clinical trial outcome prediction, we have adapted and included the following specialized models:

- DeepEnroll [20]: This model is tailored for encoding clinical trial eligibility criteria, featuring a hierarchical embedding network for disease ontology representation and an alignment model that captures the interplay between eligibility criteria and disease attributes. The eligibility criteria embeddings are merged with molecular embeddings derived from MPNN to refine trial outcome predictions.
- COMPOSE [21]: Initially designed for patient-trial matching, this model combines convolutional neural networks with memory networks to encode eligibility criteria and diseases. The resulting embeddings are integrated with molecular embeddings from MPNN to enhance trial outcome prediction capabilities.
- HINT [6]: As the current state-of-the-art in trial outcome prediction, HINT integrates a multifaceted approach. It includes a drug molecule encoder based on MPNN, a disease ontology encoder using GRAM, an eligibility criteria encoder leveraging BERT, a pharmacokinetic encoder for drug molecules, and a graph neural network to facilitate interaction between features. The composite embeddings are then processed by a prediction model to forecast trial outcomes.
- llm4top-zeroshot: This is the vanilla Qwen3 prediction framework utilizing prompt engineering strategies to guide the model’s clinical trial outcome predictions, where the base model’s inherent knowledge is directly leveraged without task-specific fine-tuning. This approach contrasts with our proposed LoRA-enhanced version that adapts the model through parameter-efficient domain adaptation.

In general, clinical trials are usually carried out in three phases: phase I examines the toxicity and side effects of the drug, phase II assesses the drug’s efficacy (that is, whether it works), and phase III focuses on determining the effectiveness of the drug in comparison with current standard practices. The experimental results underscore the efficacy of employing the llm4top for predicting clinical trial outcomes, demonstrating superior accuracy and precision when compared to conventional machine learning methodologies in from Tables 1, 2, 3.

**Table 1.** Trial outcome prediction results compared between llm4top and baselines for phase I trials.

| Phase I Trials<br># train: 1,044; # valid: 116; # test: 627; # patients per trial: 45 |  |  |  |
| --- | --- | --- | --- |
| Method | PR-AUC | F1 | ROC-AUC |
| LR [18] | 0.500 ± 0.005 | 0.604 ± 0.005 | 0.520 ± 0.006 |
| RF [18] | 0.518 ± 0.005 | 0.621 ± 0.005 | 0.525 ± 0.006 |
| XGBoost [18] | 0.513 ± 0.06 | 0.621 ± 0.007 | 0.518 ± 0.006 |
| AdaBoost [18] | 0.519 ± 0.005 | 0.622 ± 0.007 | 0.526 ± 0.006 |
| kNN+RF [18] | 0.531 ± 0.006 | 0.625 ± 0.007 | 0.538 ± 0.005 |
| FFNN [19] | 0.547 ± 0.010 | 0.634 ± 0.015 | 0.550 ± 0.010 |
| DeepEnroll [20] | 0.568 ± 0.007 | 0.648 ± 0.011 | 0.575 ± 0.013 |
| COMPOSE [21] | 0.564 ± 0.007 | 0.658 ± 0.009 | 0.571 ± 0.011 |
| HINT [6] | 0.567 ± 0.010 | 0.665 ± 0.010 | 0.576 ± 0.008 |
| llm4top-zeroshot | 0.553 ± 0.020 | 0.711 ± 0.017 | 0.502 ± 0.021 |
| llm4top-lora | **0.616 ± 0.024** | **0.711 ± 0.016** | **0.616 ± 0.020** |

**Table 2.** Trial outcome prediction results compared between llm4top and baselines for phase II trials.

| Phase II Trials<br># train: 4,004; # valid: 445; # test: 1,653; # patients per trial: 183 |  |  |  |
| --- | --- | --- | --- |
| Method | PR-AUC | F1 | ROC-AUC |
| LR [18] | 0.565 ± 0.005 | 0.555 ± 0.006 | 0.587 ± 0.009 |
| RF [18] | 0.578 ± 0.008 | 0.563 ± 0.009 | 0.588 ± 0.009 |
| XGBoost [18] | 0.586 ± 0.006 | 0.570 ± 0.009 | 0.600 ± 0.007 |
| AdaBoost [18] | 0.586 ± 0.009 | 0.583 ± 0.008 | 0.603 ± 0.007 |
| kNN+RF [18] | 0.594 ± 0.008 | 0.590 ± 0.006 | 0.597 ± 0.008 |
| FFNN [19] | 0.604 ± 0.010 | 0.599 ± 0.012 | 0.611 ± 0.011 |
| Deep Enroll [20] | 0.600 ± 0.010 | 0.598 ± 0.007 | 0.625 ± 0.008 |
| COMPOSE [21] | 0.604 ± 0.007 | 0.597 ± 0.006 | 0.628 ± 0.009 |
| HINT [6] | 0.629 ± 0.009 | 0.620 ± 0.008 | 0.645 ± 0.006 |
| llm4top-zeroshot | 0.554 ± 0.020 | **0.713 ± 0.013** | 0.500 ± 0.001 |
| llm4top-lora | **0.652 ± 0.015** | 0.700 ± 0.011 | **0.655 ± 0.001** |

**Table 3.** Trial outcome prediction results compared between llm4top and baselines for phase III trials.

| Phase III Trials<br># train: 3,092; # valid: 344; # test: 1,140; # patients per trial: 1,418 |  |  |  |
| --- | --- | --- | --- |
| Method | PR-AUC | F1 | ROC-AUC |
| LR [18] | 0.687 ± 0.005 | 0.698 ± 0.005 | 0.650 ± 0.007 |
| RF [18] | 0.692 ± 0.004 | 0.686 ± 0.010 | 0.663 ± 0.007 |
| XGBoost [18] | 0.697 ± 0.007 | 0.696 ± 0.005 | 0.667 ± 0.005 |
| AdaBoost [18] | 0.701 ± 0.005 | 0.695 ± 0.005 | 0.670 ± 0.004 |
| kNN+RF [18] | 0.707 ± 0.007 | 0.698 ± 0.008 | 0.678 ± 0.010 |
| FFNN [19] | 0.747 ± 0.011 | 0.748 ± 0.009 | 0.681 ± 0.008 |
| DeepEnroll [20] | 0.777 ± 0.008 | 0.786 ± 0.007 | 0.699 ± 0.008 |
| COMPOSE [21] | 0.782 ± 0.008 | 0.792 ± 0.007 | 0.700 ± 0.007 |
| HINT [6] | 0.811 ± 0.007 | 0.847 ± 0.009 | **0.723 ± 0.006** |
| llm4top-zeroshot | 0.7467 ± 0.012 | 0.855 ± 0.001 | 0.502 ± 0.002 |
| llm4top-lora | **0.8321 ± 0.010** | **0.856 ± 0.007** | 0.693 ± 0.015 |

## VI. DISCUSSION

The experimental findings validate that utilizing llm4top for clinical trial outcome prediction enhances accuracy and precision, outperforming traditional machine learning techniques. These results empirically demonstrate that processing modals like drugs, diseases, and trial eligibility criteria as text inputs into llm4top yields superior outcomes compared to designing and integrating separate models for each modality. This streamlined approach not only simplifies the modeling process but also boosts overall predictive performance. The study reveals that llm4top effectively discerns intricate patterns and correlations within clinical trial data when supervised finetuned training is applied, although it performs less effectively with zero-shot prompts, which is understandable given the absence of clinical data in the base model training. However, it’s acknowledged that the methodology may not be universally applicable to all categories of clinical trials or datasets. Future research should explore the adaptability of this technique across a diverse range of datasets and scenarios, thereby reinforcing its validity and expanding its applicability.

## V. CONCLUSION

In this paper, the potential of llm4top in predicting clinical trial outcomes is demonstrated. By reformulating the prediction task as a binary classification problem, the capabilities of large language models are effectively utilized to enhance the predictive accuracy of clinical trial outcomes. As the first framework with GUI developed to cover the entire trial outcome prediction pipeline, it dramatically reduce the process of deploying the AI model. The findings presented herein offer a promising new approach for improving clinical trial outcomes prediction, with significant implications for research, drug development, and patient care. However, the methodology still has limitations. Foremost among these is its reliance on the accessibility and thoroughness of historical clinical trial data for training, which may not always be available or comprehensive. Moreover, the assumption that trial outcomes can be accurately captured within a binary classification framework may not be universally applicable. Future research should address these constraints, perhaps by developing more nuanced predictive models capable of handling multi-class or continuous outcomes. Furthermore, the incorporation of these models into operational clinical software systems encounters legal and regulatory obstacles, since these models need to be approved as medical devices.

## Data Availability

All data produced are available online at https://clinicaltrials.gov/

## REFERENCES

[1] Yang, An, et al. “Qwen3 technical report.” arXiv preprint 2505.09388 (2025).

[2] Benjamin E Blass. 2015. Basic principles of drug discovery and development. Elsevier.

[3] Kaitlyn M Gayvert, Neel S Madhukar, and Olivier Elemento. 2016. A data-driven approach to predicting successes and failures of clinical trials. Cell chemical biology 23, 10 (2016), 1294–1301.

[4] Youran Qi and Qi Tang. 2019. Predicting phase 3 clinical trial results by modeling phase 2 clinical trial subject level data using deep learning. In Machine Learning for Healthcare Conference. PMLR, 288–303.

[5] Andrew W Lo, Kien Wei Siah, and Chi Heem Wong. 2019. Machine learning with statistical imputation for predicting drug approval. Harvard Data Science Review (June 2019).

[6] Tianfan Fu, Kexin Huang, Cao Xiao, Lucas M Glass, and Jimeng Sun. 2022. HINT: Hierarchical interaction network for clinical-trial-outcome predictions. Patterns 3, 4 (2022), 100445.

[7] Devlin, Jacob, et al. “Bert: Pre-training of deep bidirectional transformers for language understanding.” Proceedings of the 2019 conference of the North American chapter of the association for computational linguistics: human language technologies, volume 1 (long and short papers). 2019.

[8] Zifeng Wang and Jimeng Sun. 2022. Trial2vec: Zero-shot clinical trial document similarity search using self-supervision. arXiv preprint 2206.14719.

[9] Reinisch, M., He, J., Liao, C., Siddiqui, S.A., Xiao, B.: Ctp-llm: Clinical trial phase transition prediction using large language models. arXiv preprint 2408.10995 (2024)

[10] Lai, H., Ge, L., Sun, M., Pan, B., Huang, J., Hou, L., Yang, Q., Liu, J., Liu, J., Ye, Z., et al.: Assessing the risk of bias in randomized clinical trials with large language models. JAMA Network Open 7(5), 2412687– 2412687 (2024)

[11] Jin, Q., Wang, Z., Floudas, C.S., Chen, F., Gong, C., Bracken-Clarke, D., Xue, E., Yang, Y., Sun, J., Lu, Z.: Matching patients to clinical trials with large language models. ArXiv (2023)

[12] Markey, N., El-Mansouri, I., Rensonnet, G., Langen, C., Meier, C.: From rags to riches: Using large language models to write documents for clinical trials. arXiv preprint 2402.16406 (2024)

[13] OpenAI: GPT-4o Mini: Advancing Cost-Efficient Intelligence. https://openai.com/index/gpt-4o-mini-advancing-cost-efficient-intelligence/. Accessed: 202410-26

[14] OpenAI: GPT-3.5. https://chatgpt.com/g/g-F00faAwkE-open-a-i-gpt-3-5. Accessed: 2024-10-26

[15] OpenAI: Hello GPT-4. https://openai.com/index/hello-gpt-4o/. Accessed: 202410-26

[16] Dubey, A., Jauhri, A., Pandey, A., Kadian, A., Al-Dahle, A., Letman, A., Mathur, A., Schelten, A., Yang, A., Fan, A., et al.: The llama 3 herd of models. arXiv preprint 2407.21783 (2024)

[17] Jin, Shuyi, et al. “Can artificial intelligence predict clinical trial outcomes?.” arXiv preprint 2411.17595 (2024).

[18] Fan Z, Xu F, Li C, Yao L. Application of KPCA and AdaBoost algorithm in classification of functional magnetic resonance imaging of Alzheimer’s disease. Neural Comput Appl. 2020;32:5329–5338. 10.1007/s00521-02004707-y.

[19] Tranchevent L-C, Azuaje F, Rajapakse JC. A deep neural network approach to predicting clinical outcomes of neuroblastoma patients. BMC Med Genomics. 2019;12(Suppl 8):178. 10.1186/s12920-019-0628-y.

[20] Zhang X, Xiao C, Glass LM, Sun J. DeepEnroll: patient-trial matching with deep embedding and entailment prediction. arXiv. 2020. 10.48550/arXiv.2001.08179.

[21] Gao J, Xiao C, Glass LM, Sun J. COMPOSE: cross-modal pseudo-siamese network for patient trial matchingKDD ‘20: Proceedings of the 26th ACM SIGKDD International Conference on Knowledge Discovery & Data Mining:803–812. 10.1145/3394486.3403123.

[22] Hu E J, Shen Y, Wallis P, et al. Lora: Low-rank adaptation of large language models[J]. arXiv preprint 2106.09685, 2021.

[23] Gao, J., Xiao, C., Glass, L.M., and Sun, J. (2020). COMPOSE: cross-modal pseudo-siamese network for patient trial matching. In Proceedings of the 26th ACM SIGKDD International Conference on Knowledge Discovery & Data Mining, pp. 803–812.

[24] Zhang, X., Xiao, C., Glass, L.M., and Sun, J. (2020). Deepenroll: patienttrial matching with deep embedding and entailment prediction. In Proceedings of The Web Conference 2020, pp. 1029–1037.

[25] Tranchevent, L.-C., Azuaje, F., and Rajapakse, J.C. (2019). A deep neural network approach to predicting clinical outcomes of neuroblastoma patients. BMC Med. Genomics 12, x1–11.

[26] F. Pedregosa, G. Varoquaux, A. Gramfort, V. Michel, B. Thirion, O. Grisel, M. Blondel, P. Prettenhofer, R. Weiss, V. Dubourg, J. Vanderplas, A. Passos, D. Cournapeau, M. Brucher, M. Perrot, and E. Duchesnay. 2011. Scikit-learn: Machine Learning in Python. Journal of Machine Learning Research 12 (2011), 2825–2830.

